# Management of rheumatic diseases in the times of COVID-19 pandemic- perspectives of rheumatology practitioners from India

**DOI:** 10.1101/2020.04.03.20048389

**Authors:** Latika Gupta, Durga Prasanna Misra, Vishwesh Agarwal, Suma Balan, Vikas Agarwal

## Abstract

**Objective:** The Coronavirus disease 19 (COVID-19) pandemic has led to widespread concerns about the risk of infection in patients with rheumatic diseases (RD) receiving disease modifying ant-rheumatic drugs (DMARDs) and other immunosuppressants (IS).

**Methods:** A SurveyMonkey® based electronic survey was conducted amongst members of the Indian Rheumatology Association to understand the need for changes in prevailing practices.

**Results:** Of the 861 invitees, 221 responded. In the wake of the pandemic, 47.5% would reduce biological DMARDs (bDMARDs) while only 12.2% would reduce the use of conventional synthetic DMARDs. 64.2% were likely to defer change in IS, the reluctance being most with rituximab (58.3%) followed by cyclophosphamide (53.3%), anti-tumor necrosis factor alpha agents (52.4%) and Janus kinase inhibitors (34.39%).

Hydroxychloroquine was the preferred choice (81.9%) for the treatment of COVID-19 followed by protease inhibitors (22.1%) and intravenous immunoglobulin (8.1%). Chloroquine was less preferred (19%). More than two-thirds (70.5%) believed that COVID-19 might trigger macrophage activation syndrome. Social distancing (98.1%) and hand hygiene (74.6%) were recommended by majority. 62.8% would avoid touch for clinical examination whenever feasible.

**Conclusion:** Most rheumatologists perceived the need to change treatment of RDs during the COVID-19 pandemic; reduce immunosuppression and defer the usage of rituximab and bDMARDs.

**Key messages:** *What is already known about this subject?:* Patients with rheumatic diseases receiving glucocorticoids, disease modifying ant-rheumatic drugs and other immunosuppressants have increased susceptibility to infections including respiratory tract infections

*What does this study add?:*

○ There is an urgent need to revise the management of rheumatic diseases as perceived by a large group of practicing rheumatologists in India in the times of the COVID-19 pandemic.
○ There is reluctance to initiate biological DMARDs (especially Rituximab and anti-TNF agents), tsDMARDs (JAK inhibitors) and cyclophosphamide.
○ There is an inclination to prescribe hydroxychloroquine (HCQ) even for rheumatic diseases with weak level of evidence.

*How might this impact on clinical practice?:* This might identify areas to be addressed in a Delphi exercise to develop expert evidence to guide the management of RDs during the pandemic.

## Introduction

The worldwide Coronavirus disease 19 (COVID-19) pandemic caused by the Severe Acute Respiratory Syndrome Coronavirus 2 (SARS-CoV-2) has led to concerns about potentially greater susceptibility of patients with rheumatic diseases (RDs) who are on disease modifying antirheumatic drugs (DMARDs), and other immunosuppressants (IS).[1] With an estimated global burden of 20 million for rheumatoid arthritis alone, and much higher for other RDs combined, there is an urgent need to determine the change in current treatment strategies in the wake of the pandemic. [2]

Patients with RDs have heightened susceptibility to infections, both due to long-term IS, and the disease itself.[3] A large proportion of them are elderly, with comorbid illnesses such as hypertension, diabetes mellitus and cardiac disease, putting them at considerable risk for COVID-19 and poor outcome.[4] Thus, it is imperative to understand rheumatologists’ perspectives on managing the RDs in such a situation and guide their current and future care. A survey was designed for this purpose.

## Methods

### Design of the questionnaire

The survey featured 31 questions of the knowledge and opinion set, wherein the Likert scale was used for the latter. The specific areas covered included continuity of medical care, treatment of RDs during the pandemic, and preventive as well as therapeutic strategies for COVID-19 and prevalent fears. Content and face validity were tested by 4 rheumatologists and one medical student. The survey was drafted after 3 rounds of revision, and took 5 minutes to administer.

Participant selection: Registered members of Indian Rheumatology Association were invited over email and WhatsApp®. Eligible participants had six days to voluntarily complete the questionnaire (March 22-28 2020). Checklist for Reporting Results of Internet E-surveys was adhered to.[5]

Exemption from review was obtained from the institute ethics committee (2018-62-IP-EXP) of SGPGIMS, Lucknow as per local guidelines.[6] Descriptive statistics were used and figures downloaded from surveymonkey.com®.

## Results

### Characteristics of survey respondents

Of 861 invitees, 221 (25.7%) responded. 92.7% were treating adults predominantly while 44.7% were treating children as well; 48.4% practiced at an academic institute. Respondent characteristics are detailed in Table 1.

### Change in patient management during the COVID-19 pandemic

Most rheumatologists perceived that they would change their approach towards management patients with lupus, myositis, systemic-sclerosis and rheumatoid arthritis, more than spondyloarthritis, fibromyalgia and osteoarthritis (Table 2).

The pandemic led 47.5% respondents to reduce the use of biological DMARDs (bDMARDs), whereas only 12.2% reduced the use of conventional synthetic DMARDs (csDMARDs). 66.5% were more inclined to initiate hydroxychloroquine in patients with borderline indications, while 14% disagreed with this approach (Figure 1). An earlier taper of glucocorticoids was preferred by 57.9% respondents in inactive disease. Nearly two-thirds (64.2%) were less likely to change the major IS in a patient with impending flare, with 58.3% deferring a switch to Rituximab (RTX) followed closely by cyclophosphamide (CYC), anti-TNFs and Janus Kinase inhibitors (JAKinibs) and other bDMARDs (Figure 1F).

**Figure 1.**
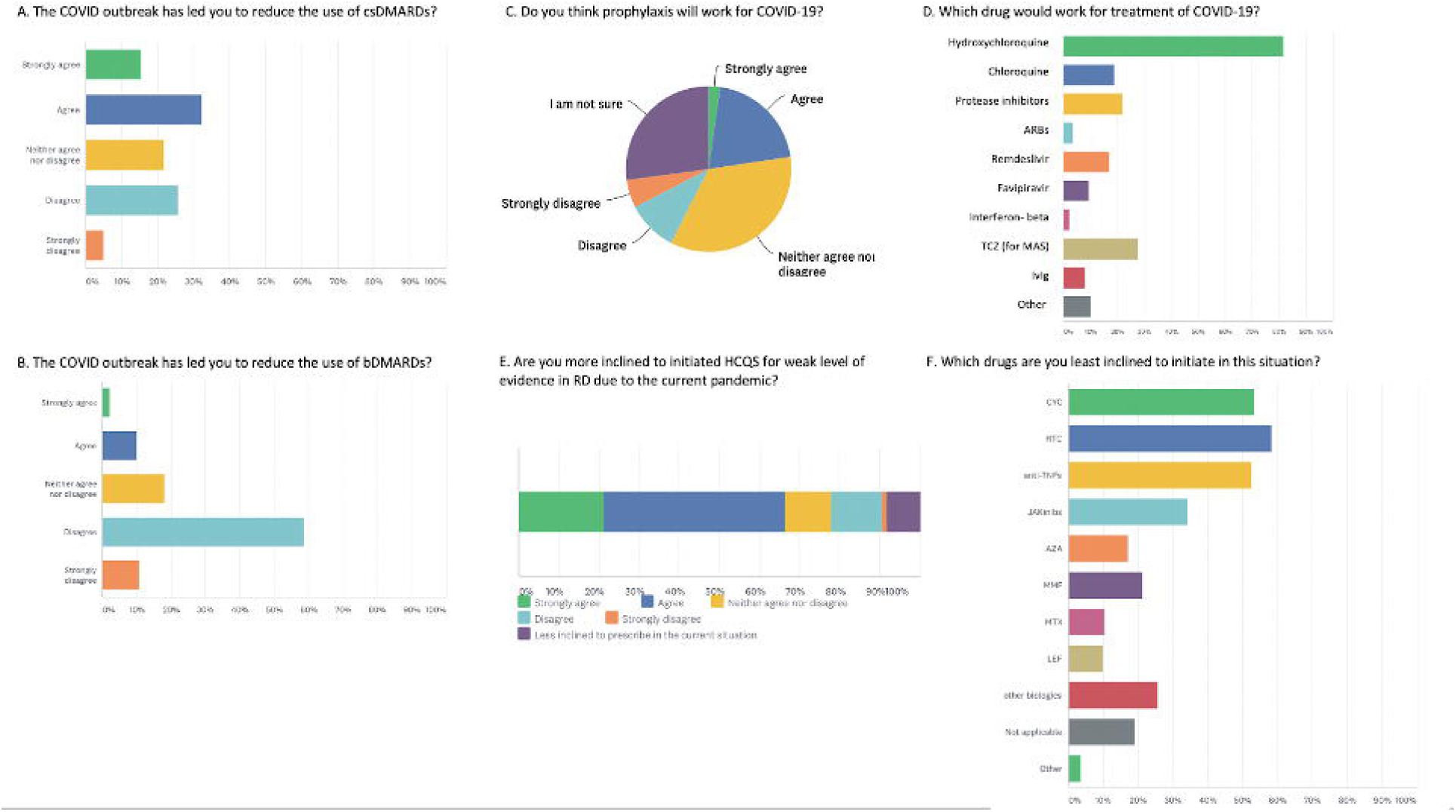
Opinion of rheumatologists on the change in management of rheumatic diseases in the times of the COVID-19 pandemic

For patients in remission and due for IS infusions, 29.8% would defer these infusions while 20.8% would not, and another 39.3% and 20.3% would decide based on severity of the primary disease or the drug being used, respectively. 61% were avoiding Ibuprofen, and 37.10% advised vaccination against other viruses more often during the pandemic. 48.4% rheumatologists had deferred the routine workup for co-morbidities and one-third would consider avoiding the use of ACEi/ARBs in RDs, while another one-third were unsure regarding this. Almost three-fourth (70.5%) respondents felt that COVID-19 could cause macrophage activation syndrome (MAS) and Tocilizumab was the preferred drug for treatment (27.6%).

### Change in delivery of patient care during the pandemic

Due to disrupted medical services, 60.2% resorted to virtual consulting, of which roughly one-thirds (30.7%) used WhatsApp®, while 16.2% and 13.12% did so over email and video calls respectively. Only 19% of respondents were continuing their clinics at the time of the survey.

### Treatment and prophylaxis of COVID-19

Hydroxychloroquine (HCQ) was the preferred choice for treatment (81.9% respondents) followed by protease inhibitors (22.17%) and IVIG (8.14%). Chloroquine was less popular (19%). 22.6% felt that HCQ prophylaxis might work for COVID-19, and the same number prescribed prophylaxis to their patients, while 27.15% were not sure if it would be beneficial. The rest 50.2% did not feel the need of HCQ prophylaxis in RDs.

### COVID-19 specific preventive measures

Almost all recommended social distancing followed by hand hygiene directives by fewer though mask usage were much lower (Table 2). Nearly two-thirds would avoid touch for a clinical examination in patients who felt fine and were likely to be in remission.

### Prevalent concerns pertaining to COVID-19

Among respondents, the most prevalent fears were passing on the infection to their family followed closely by patients getting infected during travels, and respondents getting infected themselves. Most felt the pandemic would last 3-6 months and suggested various measures to reduce the potential community transmission of COVID-19 (Table 2).

## Discussion

Due to the COVID-19 pandemic, rheumatologists were hesitant in the usage of bDMARDs (especially RTX followed by anti-TNFs), targeted synthetic (tsDMARDs) and CYC and more inclined to prescribe HCQ for treatment of RDs whereas csDMARDs were perceived as safe. There was lack of consensus on continuing IS infusions. IVIG usage was favoured by a minority however, it still merits consideration.

Reasonable risk of viral activation has previously been described with RTX and JAKinibs, and thus the hesitation in prescribing various bDMARDs as well as JAKinibs is not unfounded. However, currently, no data are available on the specific risk of respiratory viral infections due to JAKinibs. While some have advocated the use of JAKinibs to inhibit viral entry through AP-2 associated kinase 1 (AAK1) mediated endocytosis, this is likely to be successful at supra-therapeutic doses, raising significant safety concerns. [7] However, data on influenza risk with anti-TNFs is virtually non-existent. [8] D’Antiga et al suggested that Corona viruses might not preferentially affect immunosuppressed post-transplant patients, however, patients with RDs are likely to have heightened risk, as has been seen in the elderly and those with other co-morbid cardiac or lung disease.[9]

Disease flares can potentially be induced by the COVID-19, as seen in RDs by most endogenous retroviruses as well as acquired viral infections.[8] Previously higher risk of infections has been reported in those with higher disease activity, and vice versa. [8] While most rheumatologists believed that COVID-19 may trigger MAS, it might be difficult to distinguish cytopenia and hyperferritinemia due to increased disease activity. The consensus was on the use of Tocilizumab in MAS, possibly backed by a case series, which remains to be confirmed in ongoing trials. [10] The feasibility of screening for SARS-CoV-2 before initiation of bDMARDs needs to be explored as previously suggested.[11]

There was unanimous agreement on use of HCQ for treatment of COVID-19, with more than two-thirds weighing towards initiating it in patients with otherwise low evidence disease-specific indications. This may be attributed to less toxic nature, fewer interactions, a wider therapeutic index, greater in-vitro efficacy against the COVID-19 and years of experience of rheumatologists.[12] A prominent national regulatory body has issued an advisory favouring prophylaxis with hydroxychloroquine for healthcare workers and close contacts of patients with COVID-19[13] However, caution is needed as reports of toxicity have emerged with the use of prophylaxis.[14] The usage of ACEi/ARBs was debateable; there exists little evidence to recommend continuation or discontinuation of either drug.[15]

In times of widespread travel bans and jeopardised medical services, there is a felt need for shift to virtual consulting. India recently legalised teleconsultations to the same effect. [16] A pandemic of such magnitude is not without psychologic impact on the treating physicians either. The dominant fears in this situation were mostly related to transmission to family and the patients. The management of the disease was most likely to be affected in the CTD spectrum of RDs, suggesting the need to develop evidence for a triage-in-rheumatology protocol bracing for the times ahead.

A strength of our survey was that 60% of the respondents had been in rheumatology practice for more than 5 years. Mhaskar et al. have reported 73% concordance between decision analysis driven by expert consensus with evidence gathered from RCTs.[17] Considering the potential limitations of generating evidence in the face of a global crises, it might be imperative to embark on a Delphi exercise to generate expert-opinion, while data from COVID-19 rheum registries in progress is awaited.[18]

The present survey provides perspectives on rheumatology care during the COVID-19 pandemic from a single country and the response rate was modest, albeit represents the viewpoint of a large number of rheumatologists. The findings of the survey may help shape future evidence-based opinions on managing patients with IS during COVID-19 pandemic.

## Conclusion

Most practicing rheumatologists in India are of the opinion that the current management of RDs need change during the COVID-19 pandemic with a greater caution in usage of bDMARDs. Consensus favoured usage of HCQ for treatment of COVID-19 though its role in prophylaxis remains unresolved.

## Data Availability

All data are available for review

---

Table 1. Respondent characteristics

Table 2. Responses to the questionnaire

Supplementary Table 1. Survey questions.

## Competing interests

The authors declare that there is no conflict of interest

## Contributorship

All authors were involved in ideation, data collection and manuscript preparation. All authors agree with the submitted version of the manuscript, take responsibility for the content of the entire manuscript, and affirm that any queries related to any aspect of the same are appropriately managed.

## Acknowledgements

None

## Funding statement

This study was not funded.

## Ethical approval information

Exemption from review was obtained from the institute ethics committee (2018-62-IP-EXP) of SGPGIMS, Lucknow as per local guidelines

## Data sharing statement

All data pertaining to the study is included in the manuscript and as supplementary material.

## Patient and Public Involvement

The survey was completely anonymised and informed consent taken from the respondents at the beginning of the exercise.

## Notes

### Competing Interest Statement

The authors have declared no competing interest.

### Funding Statement

Not applicable

